# EMERGE: Evaluating the value of measuring random plasma glucose values for managing hyperglycemia in the inpatient setting

**DOI:** 10.1101/2022.08.23.22279142

**Authors:** Saba Manzoor, Mike Colacci, Jason Moggridge, Michelle Gyenes, Tor Biering-Sørensen, Mats C. Højbjerg Lassen, Fahad Razak, Amol Verma, Shohinee Sarma, Michael Fralick

## Abstract

**Importance:** A diagnosis of diabetes is considered when a patient has hyperglycemia with a random plasma glucose ≥200 mg/dL. However, in the inpatient setting, hyperglycemia is frequently non-specific, especially among patients who are acutely unwell. As a result, patients with transient hyperglycemia may be incorrectly labeled as having diabetes, leading to unnecessary treatment, and potential harm.

**Design, Setting, and Participants:** We conducted a multicentre cohort study of patients hospitalized at seven hospitals in Ontario, Canada and identified those with a glucose value ≥200 mg/dL. We validated a definition for diabetes using manual chart review that included physician notes, pharmacy notes, home medications, and hemoglobin A1C. Among patients with a glucose value ≥200 mg/dL, we identified patients without diabetes who received a diabetes medication, and the number who experienced hypoglycaemia during the same admission.

**Main Outcomes and Measures:** To determine the diagnostic value of using random blood glucose to diagnose diabetes in the inpatient setting, and its impact on patient outcomes.

**Results:** We identified 328,786 hospitalizations from hospital between 2010 and 2020. A blood glucose value of ≥200 mg/dL had a positive predictive value of 68% and a negative predictive value of 90% for a diagnosis of diabetes. Of the 76,967 patients with an elevated glucose value reported, 16,787 (21.8%) did not have diabetes, and of these, 5,375 (32%) received a diabetes medication. Hypoglycemia was frequently reported among the 5,375 patients that received a diabetes medication, with 1,406 (26.2%) experiencing hypoglycemia and 405 (7.5%) experiencing severe hypoglycemia.

**Conclusions and Relevance:** Elevated plasma glucose in hospital is common but does not necessarily indicate a patient has diabetes. Furthermore, it can lead to treatment with diabetes medications with potential harm. Our findings highlight that clinicians should be cautious when responding to elevated random plasma glucose tests in the inpatient setting.

## Introduction

Diabetes mellitus continues to be a leading cause of preventable morbidity worldwide, highlighting the need for effective diagnostic testing.^1^ A diagnosis of diabetes is established based on symptoms of hyperglycemia accompanied by an elevated glycated hemoglobin (HbA1c) ≥6.5%, a fasting plasma glucose ≥ 126 mg/dl, a 2-hour plasma glucose ≥ 200 mg/dl in a 75g oral glucose tolerance test or a random plasma glucose ≥ 200 mg/dl. In asymptomatic patients, confirmatory testing is required to establish a diagnosis.^2,3^ Despite the clarity of these guidelines, the subtleties of needing confirmatory testing can be missed, particularly in the inpatient setting where patients may experience hyperglycemia due to acute illness.^4^ For these reasons, inpatients with hyperglycemia may be incorrectly labelled as having diabetes based on isolated results from random plasma glucose testing.

Another limitation of using the random plasma glucose test to diagnose diabetes is its Grade D evidence rating, based on limited evidence to support the guideline recommendation.^2,5^ For this reason, asymptomatic patients with an elevated random plasma glucose require confirmatory testing with another diagnostic test (e.g., fasting plasma glucose, HbA1c). Conversely, the fasting plasma glucose and HbA1c tests have a stronger Grade B evidence rating, and elevated values can be confirmed with the same type of test (i.e., a HbA1c ≥6.5% in an asymptomatic patient can be confirmed by a repeat HbA1c ≥6.5%).^2^ The nuance of conducting confirmatory testing for the random plasma glucose test in asymptomatic patients is amplified in the inpatient setting, where patients are acutely unwell and may have symptoms mimicking diabetes (e.g., polydipsia) that are due to an alternate etiology.

Considering the challenges of applying the random plasma glucose test in the inpatient setting, patients without diabetes may frequently be treated for hyperglycemia in the absence of a diagnosis of diabetes. This may lead to downstream management with diabetes drugs, putting patients at increased risk of side-effects related to the medication (e.g., hypoglycemia). Hypoglycemia can be particularly concerning and life-threatening, if severe.^6^ The objective of our study was to assess the accuracy of the inpatient random plasma glucose test for a diagnosis of diabetes and its impact on patient outcomes.

## Methods

### Study Design

We conducted a cohort study of adults (≥18 years old) admitted to the inpatient medicine service in 7 hospitals (5 academic health science centres and 2 community-based teaching hospitals) in Ontario, Canada between 2010 and 2020. From 2010 to 2015, data was only available for patients admitted to, or discharged from the inpatient general internal or hospitalist medicine service. In 2016, additional data for patients admitted to, or discharged from the medical-surgical ICU was also included. The hospitals included in this study were part of the General Medicine Inpatient Initiative (GEMINI) database, as reported previously.^7^ These sites are all linked with the University of Toronto and are located in the Greater Toronto Area, the most populous metropolitan area in Canada, with a population of 7.2 million.^8^ Our study was approved by the research ethics board at the hospitals included in our study.

### Cohort Definition and Follow-Up

We included all consecutive adult patients admitted to or discharged from the inpatient medical service or medical-surgical ICU. Patients without a glucose value drawn during their hospitalization were excluded. The glucose values used to define our patient cohort were established using glucose point of care tests as well as plasma glucose testing from routine blood work. We further identified the number of patients with an elevated random blood glucose ≥200 mg/dL. The cut-off value of 200 mg/dL was selected based on international guidelines for diagnosing diabetes in hyperglycemic patients. ^2,3^ Patients were followed from the date of hospitalization to hospital discharge or in-hospital death.

### Data Sources

Administrative and clinical data were extracted from each patient’s electronic medical record. Patients’ electronic medical records were linked to data reported to the Discharge Abstract Database (DAD) and the National Ambulatory Care Reporting System (NACRS). These administrative datasets included demographic information, comorbid conditions, length of stay, and most responsible discharge diagnosis. In addition to the administrative data, clinical data including laboratory results (biochemistry, hematology), imaging results, vital signs, physician orders and medications administered to patients during their hospitalization are all included within GEMINI. The most responsible discharge diagnosis was manually coded by hospital-based chart abstractors using the International Statistical Classification of Disease and Related Health Problems (ICD-10-CA) and the Canadian Classification of Health Interventions (CCI). The accuracy of data in the GEMINI database, relative to chart review by a trained chart abstractor have previously been evaluated and all tested data fields have an accuracy of above 98%.^7,9^

### Assessment of Random Blood Glucose Test Characteristics

In addition to the administrative and clinical data automatically captured from the GEMINI database, charts were also manually reviewed by individual abstractors to extract detailed information about the patient’s course in hospital. We performed a chart review of 700 randomly selected charts, including 350 with a glucose ≥ 200 mg/dL, and 350 without a glucose ≥ 200 mg/dL. The chart review allowed us to calculate the test characteristics for not only hyperglycemia but also the ICD-10 codes for diabetes. The admission note, discharge summary and medication reconciliation note were manually reviewed to determine the test characteristics and concordance between hyperglycemia and diabetic status. At the included hospitals, the admission and discharge note are written by a physician and the medication reconciliation note is written by the pharmacist. Based on the chart review, we considered a patient to have diabetes if at least one of the following criteria were met: (i) their admission note, discharge summary, or medication reconciliation indicated the patient had diabetes, (ii) they were on a medication for diabetes prior to their hospitalization (Appendix), or (iii) a recorded hemoglobin A1c value ≥ 6.5%. We did not include discharge medications in the definition because some patients may have their diabetes medications stopped at the time of discharge and because these medications may have been inappropriately started in response to hyperglycemia.

### Study Outcomes

Our main outcome was to estimate the proportion of patients with a glucose ≥200 mg/dL who did not have diabetes (using our aforementioned definition) and received medication for diabetes. We calculated the proportion of patients who had a random plasma glucose ≥200 mg/dL but who did and did not have a diagnosis of diabetes through a multi-step approach. First, if the patient had a hemoglobin A1c ≥ 6.5% on their current admission, they were considered to have diabetes. For patients without an A1c measured, we used ICD-10 codes from a current or prior hospitalization to identify if they had diabetes. For patients with an A1c less than 6.5%, they were classified as having diabetes only if they had an ICD-10 code for diabetes from a current or prior hospitalization. Doing so allowed us not to “miss” patients who had well-controlled diabetes (i.e., A1c less than 6.5%).

For patients with a glucose ≥200 mg/dL who did not have diabetes, we assessed how often they received a medication for diabetes and how often they developed hypoglycemia. Hypoglycemia was defined as a glucose value 70 mg/dL, severe hypoglycemia was defined as a glucose value ≤ 45 mg/dL.

### Statistical Analysis

We calculated sensitivity, specificity, positive predictive value (PPV), negative predictive value (NPV), for an elevated blood glucose ≥ 200 mg/dL, where the gold standard was chart review (i.e., admission note, discharge note, medication reconciliation) determination of whether the patient had diabetes. We also calculated sensitivity, specificity, PPV, NPV, for the ICD-10 codes, where the gold standard was chart review determination of whether the patient had diabetes.

We calculated descriptive statistics to compare the baseline characteristics of patients with and without inpatient hyperglycemia. We also calculated the number and percentage of patients who did not have diabetes, received a diabetes medication and experienced hypoglycemia or severe hypoglycemia.

## Results

### Study Cohort

Between 2010 and 2020, there were 328,786 hospitalizations. We excluded 10,921 (3.3%) because there was no recorded blood glucose value. Among the remaining 317,865 patients, the mean age was 70 years, 154,313 (48.5%) were female, 5,592 (1.8%) had a diagnostic code for type 1 diabetes and 89,466 (28.1%) had a diagnostic code for type 2 diabetes (Table 1). The median hemoglobin A1c was 6.2% (IQR 5.6-7.6), glucose was 6.6 (IQR 5.6-8.6) mmol/L, creatinine was 79 (IQR 63-108) µmol/L and **C-**reactive protein (CRP) was 32.5 (IQR 6.9-102) mg/L. Within the total cohort of 317,865 hospitalizations in-hospital medications ordered included: metformin [N=48,255 (15.2%)], insulin [N= 24,993 (7.9%)], DPP4 inhibitors [N=25,701 (8.1%)], sulfonylureas [N=22,822 (7.2%)] and SGLT2 inhibitors [N=2,363 (0.7%)].

**Table 1.**
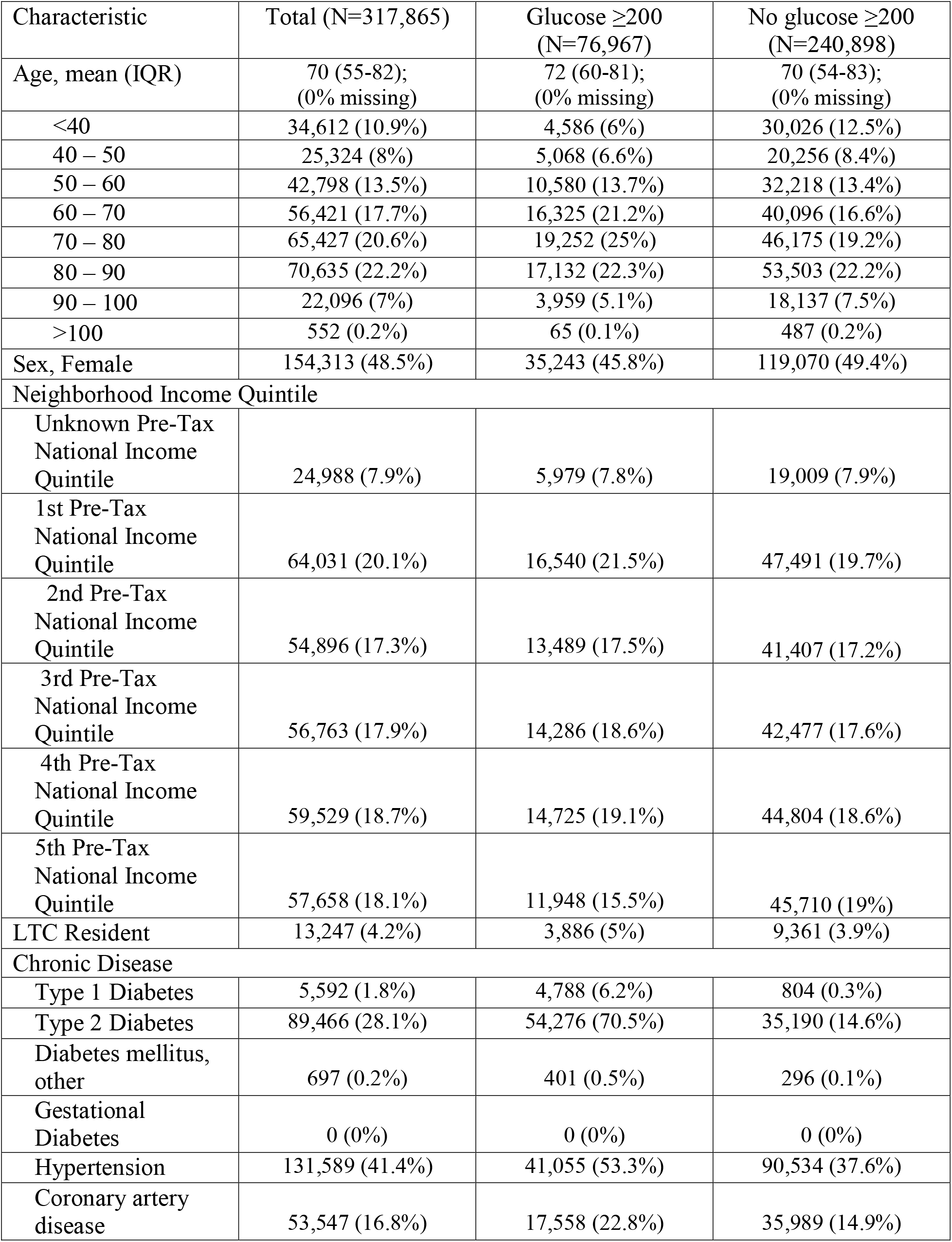
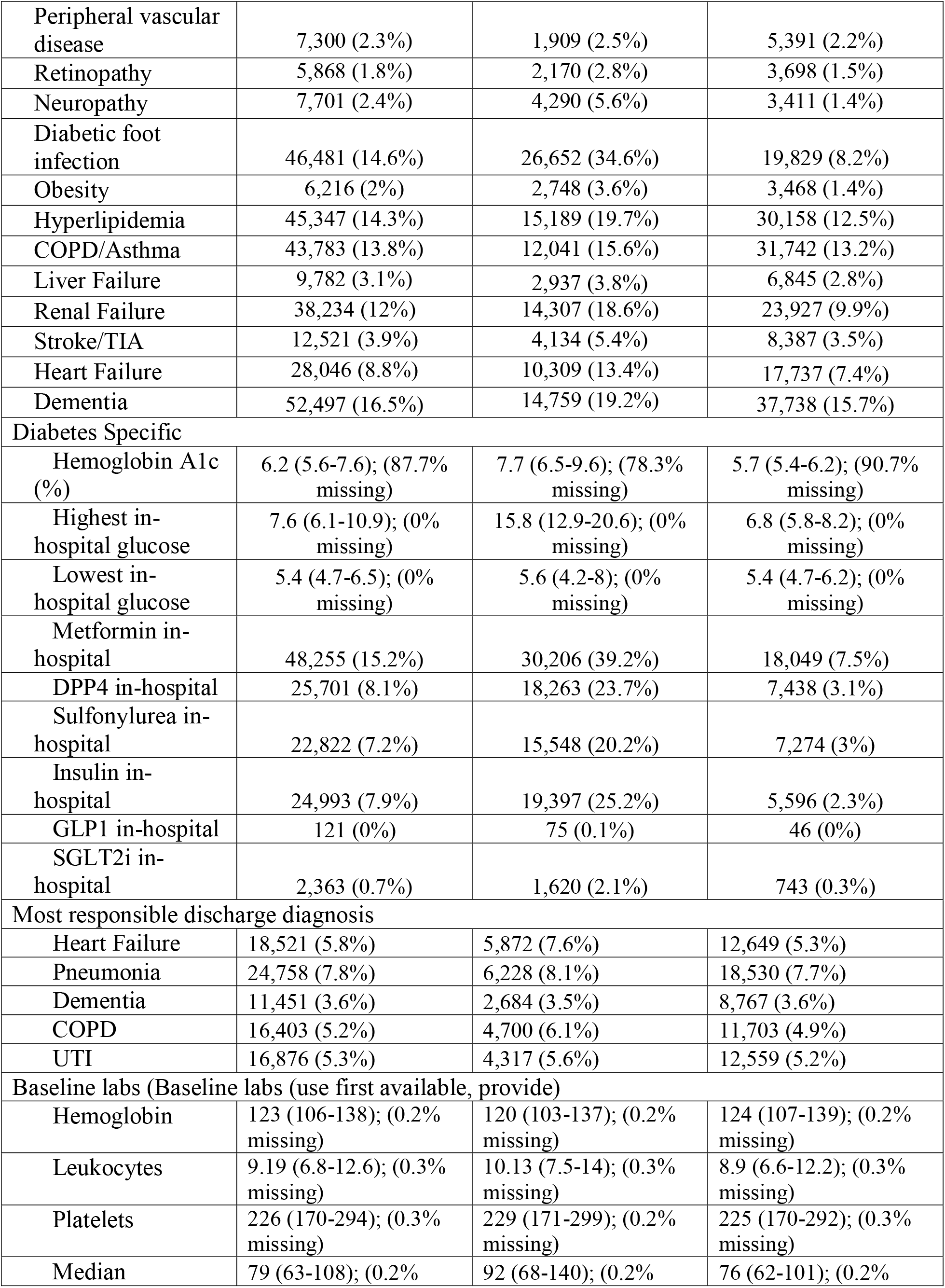
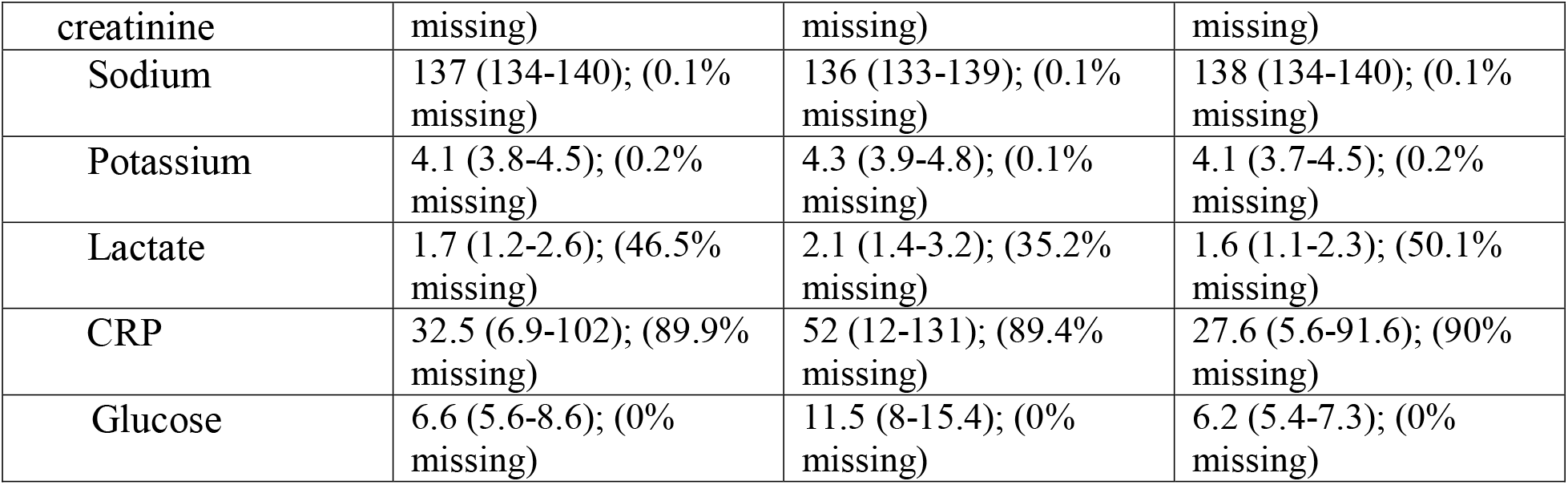
Demographic and Clinical Characteristics of the Study Population.

Of the 317,865 (96.6%) patients with a glucose value reported, 76,967 (24.2%) had at least one value ≥200 mg/dL and 240,898 (75.8%) had no glucose value ≥200 mg/dL. Patients who had blood glucose ≥ 200 mg/dl, were more likely to be male, and at higher cardiac risk (e.g., higher prevalence of hypertension, and hyperlipidemia). Patients with hyperglycemia also had a higher median hemoglobin A1c and serum glucose, a higher baseline creatinine and CRP level and were more likely to have received a diabetes medication in hospital (Table 1). The median hemoglobin A1c value for patients with at least one glucose ≥200 mg/dL was 7.7% (IQR 6.5-9.6) and for those patients with no value ≥200 mg/dL was 5.7% (5.4-6.2), but hemoglobin A1C was only available for a sub-set of patients (Table 1).

### Diagnostic accuracy of the plasma glucose and ICD-10 codes for diabetes

Compared with chart review, the sensitivity and specificity for a cut-off of 200 mg/dL was 88% and 74%, respectively. This corresponded to a positive predictive value (PPV) of 68%, and negative predictive value (NPV) of 90%. Furthermore, compared to chart review, the sensitivity of using ICD-10 codes (E10, E11, E13, and E14) to identify patients with diabetes was 97%, the specificity was 97%, the PPV was 96% and NPV was 98%. (Table 2)

**Table 2.** Diagnostic Value of Random Plasma Glucose Values based.

|  | % (95% CI) |  |
| --- | --- | --- |
| | Elevated glucose $\geq 200$ mg/dL | ICD-10 code for diabetes |
| Sensitivity | 0.88 (0.83-0.91) | 0.97 (0.95-0.99) |
| Specificity | 0.74 (0.69-0.78) | 0.97 (0.95-0.98) |
| Positive Predictive Value | 0.68 (0.64-0.71) | 0.96 (0.94-0.98) |
| Negative Predictive Value | 0.90 (0.87-0.93) | 0.98 (0.96-0.98) |

**Legend:** n=500 charts reviewed for ICD-10 codes, n=700 charts reviewed for elevated glucose

### Clinical management of elevated random plasma glucose

Among the 76,967 patients who had hyperglycemia, 16,787 (21.8%) did not have a diagnosis of diabetes. One-third (N=5375, 32%) of the patients without diabetes received a medication for diabetes, with insulin being the most commonly used medication (Table 3). Among the 5,375 patients without diabetes who received a diabetes drug, 1,406 (26.2%) developed hypoglycemia and 405 (7.5%) developed severe hypoglycemia during the same admission.

**Table 3.**
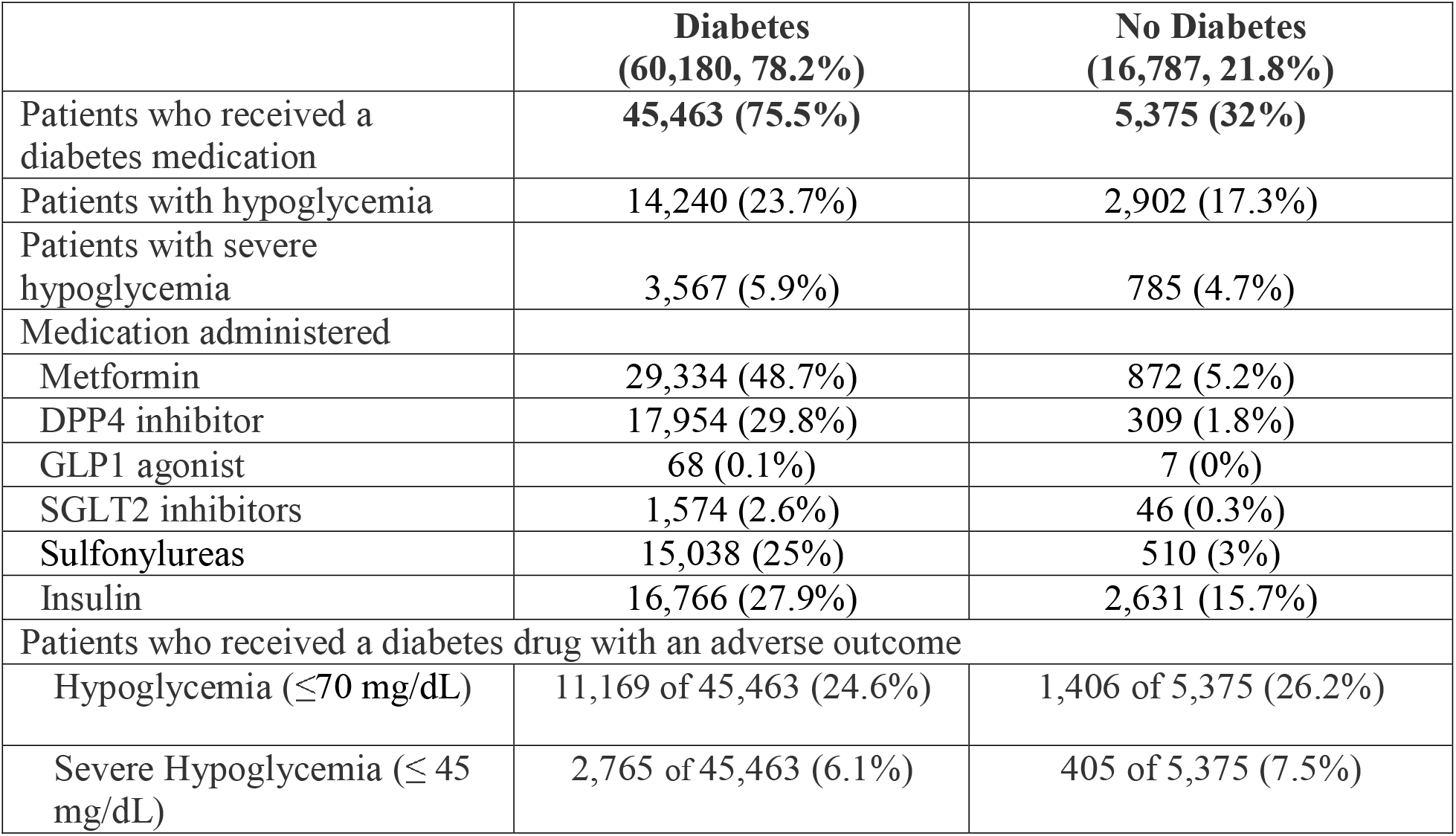
Characteristics of patients with at least one glucose ≥200 MD.

## Discussion

Our multicentre cohort study of over 300,000 hospitalized patients identified that a glucose value ≥200 mg/dL has good negative predictive value but poor positive predictive value for the diagnosis of diabetes. As a result, patients may be falsely identified as having diabetes based on a single elevated glucose measurement. Our study further demonstrated that this incorrect label is associated with receiving a diabetes medication and hypoglycemia during the same admission. These results are directly relevant to clinicians caring for patients admitted to inpatient and non-surgical ICU units in hospitals.

Previous studies have provided a wide estimate of the positive predictive value of hyperglycemia for the diagnosis of diabetes, ranging from 39% – 85%.^10–12^ The higher estimates were from studies of patients in the outpatient setting, while the 39% was from a study of inpatients similar to our study. Our study’s estimate of a positive predictive value of 68**%** confirms that hyperglycemia among inpatients does not necessarily mean a patient has diabetes. After all, hyperglycemia is a common part of the physiologic response to acute illness, regardless of whether a person has diabetes. During acute illness a cortisol and catecholamine surge can occur, leading to relative insulin deficiency, ultimately producing transient hyperglycemia that is most effectively managed by treating the underlying illness.^13^ In our study we identified that markers of inflammation (CRP) as well as illness severity (lactate) were higher among patients with hyperglycemia compared to patients who did not have hyperglycemia. This finding aligns with the known physiologic change with acute illness, and also emphasizes that inpatient hyperglycemia is often associated with an acute etiology.

Treating stress hyperglycemia with diabetes medications can potentially lead to downstream complications. Our study identified 5,375 patients without diabetes who received a diabetes medication, among whom 26.2% (n=1,406) developed hypoglycemia and 7.5% (n=405) developed severe hypoglycemia during the same admission. It is understandable why these patients received medication for diabetes, especially during a busy on-call shift, where providing insulin for a call about hyperglycemia may often be a reflexive order. Guidelines for managing hyperglycemia in the inpatient setting can also be unclear. For example, the American Diabetes Association (ADA) states that critically ill patients with a blood glucose ≥180 mg/dL should be initiated on insulin therapy, regardless of a previous diagnosis of diabetes, but the American College of Physicians (ACP) recommends against intensive insulin therapy in patients without a history of diabetes.^14^ Ultimately, the heterogeneity in clinical practice guidelines combined with a rise in inpatient volumes and complexity, can complicate clinical decision-making for managing hyperglycemia among patients that do not have diabetes.^15^

Our study has multiple limitations that should be considered. First, our diagnostic accuracy (i.e., sensitivity, specificity, PPV, NPV) for random glucose values analysis was derived from a cohort of 700 patients randomly selected for chart review, as opposed to the full cohort of 317 865 patients with a reported random plasma glucose value. Second, our study population primarily included internal medicine inpatients, which may explain why the 68% positive predictive value of the elevated blood glucose reported in our analysis was lower than the values previously reported in more heterogeneous patient populations including outpatients. Third, to identify “true” patients with diabetes in our cohort, we used ICD-10 codes. Though the sensitivity and PPV of these codes was validated at 97.5% and 96.2**%** respectively, we acknowledge this approach is still imperfect. However, other studies of inpatients have identified similar accuracy of ICD-10 codes for diabetes.^16^ Fourth, some medications for diabetes are also used for adults without diabetes (e.g., SGLT2 inhibitors) which may affect the accuracy of our definition used to identify adults with diabetes. However, during our study period GLP1 agonist use was exceedingly rare and SGLT2 inhibitors were only approved for adults with type 2 diabetes mellitus. Finally, our study did not stratify patients by diabetes type, due to the low numbers of patients with type I diabetes mellitus. However, we acknowledge the additional insight that our analysis may have offered if patients had been further divided into diabetes subtypes.

### Conclusion

In our multicentre cohort study, we found that elevated random plasma glucose values were frequently identified in inpatients without diabetes. These patients were often treated with diabetes medications which led to adverse events including severe hypoglycemia. Future research is needed to better identify which factors predispose patients with an elevated blood glucose ≥200 mg/dL to receiving treatment with diabetes-lowering medications, as these findings may potentially avert life-threatening complications in these patients.

## Data Availability

All data produced in the present study are available upon reasonable request to the authors

## Notes

**Conflicts of interest:** The authors report no relevant conflicts of interest.

### Competing Interest Statement

The authors have declared no competing interest.

### Funding Statement

This study was funded by the Banting & Best Diabetes Centre and the Sinai Department of Medicine research fund.

### Author Declarations

This research was approved through Clinical Trials Ontario (CTO) at Unity Health Toronto.

